# Robust estimation of diagnostic rate and real incidence of COVID-19 for European policymakers

**DOI:** 10.1101/2020.05.01.20087023

**Authors:** M. Català, D. Pino, M. Marchena, P. Palacios, T. Urdiales, P.J. Cardona, S. Alonso, D. López-Codina, C. Prats, E. Alvarez-Lacalle

**Affiliations:** Department of Physics. Universitat Politècnica de Catalunya (UPC-BarcelonaTech), C. Jordi Girona, 1-3, 08034 Barcelona, Spain; Comparative Medicine and Bioimage Centre of Catalonia (CMCiB), Fundació Institut d’Investigació en Ciències de la Salut Germans Trias i Pujol, Badalona, Catalonia, Spain; Experimental Tuberculosis Unit (UTE). Fundació Institut Germans Trias i Pujol (IGTP). Universitat Autònoma de Barcelona (UAB). Edifici Mar. Can Ruti Campus. Crtra. de Can Ruti, Camí de les Escoles, s/n, 08916, Badalona, Catalonia, Spain; Centro de Investigación Biomédica en Red de Enfermedades Respiratorias (CIBERES). Av. Monforte de Lemos, 3-5. Pabellón 11. Planta 0. 28029, Madrid, Spain

**Author notes:** These authors contributed equally to this work (M. Català, D. Pino, E. Alvarez-Lacalle).

## Abstract

Policymakers need a clear and fast assessment of the real spread of the epidemic of COVID-19 in each of their respective countries. Standard measures of the situation provided by the governments include reported positive cases and total deaths. While total deaths immediately indicate that countries like Italy and Spain have the worst situation as of mid April 2020, on its own, reported cases do not provide a correct picture of the situation. The reason is that different countries diagnose diversely and present very distinctive reported case fatality rate (CFR). The same levels of reported incidence and mortality might hide a very different underlying picture. Here we present a straightforward and robust estimation of the diagnostic rate in each European country. From that estimation we obtain an uniform unbiased incidence of the epidemic. The method to obtain the diagnostic rate is transparent and empiric. The key assumption of the method is that the real CFR in Europe of COVID-19 is not strongly country-dependent. We show that this number is not expected to be biased due to demography nor the way total deaths are reported. The estimation protocol has a dynamic nature, and it has been giving converging numbers for diagnostic rates in all European countries as of mid April 2020. From this diagnostic rate, policy makers can obtain an Effective Potential Growth (EPG) updated everyday providing an unbiased assessment of the countries with more potential to have an uncontrolled situation. The method developed will be used to track possible improvements on the diagnostic rate in European countries as the epidemic evolves.

## Introduction

The evolution of the epidemic in Europe has affected Spain and Italy more strongly than in other countries so far. This is clear from reported cases and fatalities in these countries [1–3]. However, comparative assessment of the spread of the pandemic in other European countries has been more difficult to assess. The reason is that the real incidence of the epidemic in each country can not be known with certainty because each country is not able to perform the same number of PCR and consequently the comparison of the ratio of those infected is difficult [4]. Policy responses have also differed, with some countries focusing tests in its clinical use in hospitals, while others have tried to use them, at least partially, to know some local chains of transmissions [5,6]. The lack of clear cross country comparison in Europe can have deep implications for the future structure of the UE since a lot of decisions are taken with a heavy influence on the sense of in-country gravity. For these reasons, it is important to have, at least, a proper measure of the relative spread of the epidemic. Policymakers must know what is the real situation in their own countries in comparison to others so that their decisions on the future of reopening and economic reconstruction are taken not from false impressions but data. In this sense, policymakers must perceive the method as unbiased, simple and robust. Most importantly, the relative comparisons between countries must be as shielded as possible from the hypothesis of the method. In this sense, methods have been developed recently [7–9] in order to asses the situation inferred from data. This work has been key to give a better picture of the situation. However, they lack the recipe-type nature needed sometimes to direct a policy response.

The focus of this paper is, thus, to introduce a method to compute the real diagnostic rate and the real incidence of COVID-19 in each European country, testing that the key hypothesis of the method is fulfilled and that, if they were to be slightly off, they would affect all countries in the same direction. In other words, we provide a recipe for policymakers that we have tested to be correct, unbiased across countries and useful to make cross-country comparison provided the evolution and prognosis of the disease in a patient is not strongly dependent on socio-economic factors and only on age, sex and previous clinical history.

We must recall here that the ability to determine the diagnostic ratio is essential to evaluate what the real number of infected people is. Knowledge of this number is not only useful to visualize the full scope of the epidemic but also to properly estimate the number of people with probable short-term immunity. In this sense, our method can be added as an empirical take of other assessments about the real incidence of the disease and to study the possibility of developing herd immunity. A large number of real infected people would be a positive scenario for policymakers while a low number will be negative. It is thus very important to err on the side of caution in all our estimates giving always the less optimistic take.

The basic structure of the paper is the following. First, we give a general overview of our framework in the methods section. Then we discuss our key assumption: the real case fatality rate (CFR) in European countries experiencing a significative incidence will be roughly the same, given the similar structure of the population. If the real CFR were to be lower, or higher, it would affect all countries in the same way and would not affect most policy decision-making since it will move all countries in the same direction. We take this real CFR to be 1% and proceed to test that, effectively, there is a strong correlation between the day of reported deaths with the number of cases taken 7-10 days before. Once a given value for the real CFR is taken, one must consider that people do not die immediately from the disease, as it takes roughly 18 days after infection [10–12]. In other words, the present values of the death toll can provide an estimation of the number of infected people 18 days ago. Knowing the number of infected people at present, not 18 days in the past, is crucial. We attack this problem considering that people who become infected are usually diagnosed a few days after the onset of the symptoms, which can be 8 to 14 days after infection occurs. By comparing the number of people diagnosed on a certain date with our estimation of the real number of infected people, we can estimate what percentage of the cases are being diagnosed. We can calculate this for different countries and regions and test how this ratio has changed dynamically as the epidemic advanced.

In the results section, we provide a full detailed description of how this fraction has become steady in the last weeks. We demonstrate that the percentage of diagnosis throughout the development of the epidemic has taken values that gradually converge for most countries. This gives a final clear picture showing the rate of diagnosis for each country. Using this rate is straightforward to give a present-day estimate of the incidence given the number of reported infected people in each country as long as we can observe that the rate of diagnosis remains fairly constant. For policymakers, we have constructed an index named Effective Potential Growth (EPG) that combines this information with the growth rate of the epidemic to provide insight regarding which countries are, comparatively and in the short-term, in the most potentially complicated situation [9].

## Methods

### Framework of our methodology

Our analysis will be applied to European countries with a minimum of 500 deaths on April 15 2020 so that we can guarantee a minimum statistical significance. The analyzed countries are: Belgium, France, Germany, Italy, Netherlands, Portugal, Spain, Sweden, Switzerlands and United Kingdom. Our two core assumptions are that the real CFR in all European countries is roughly the same and that reported data of death due COVID-19 is uniform in all European countries under consideration. We will address these two hypothesis in the following sections. With these assumptions we need to carry out four steps, as indicated in Fig. 1, to obtain the percentage of diagnosis. First, using a common reference CFR = 1% and, given the reported reported death count, we estimate the number of cases 18 days ago. According to medical reports people die between 15 and 22 days after the development of the first symptoms [13]. This time to death, TtD, after the development of the first symptoms will not be country-specific for demographic reasons. The estimated number of infected people with the disease at time *t* (see process in Fig. 1(A)) reads:

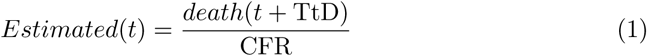

**Fig 1.**
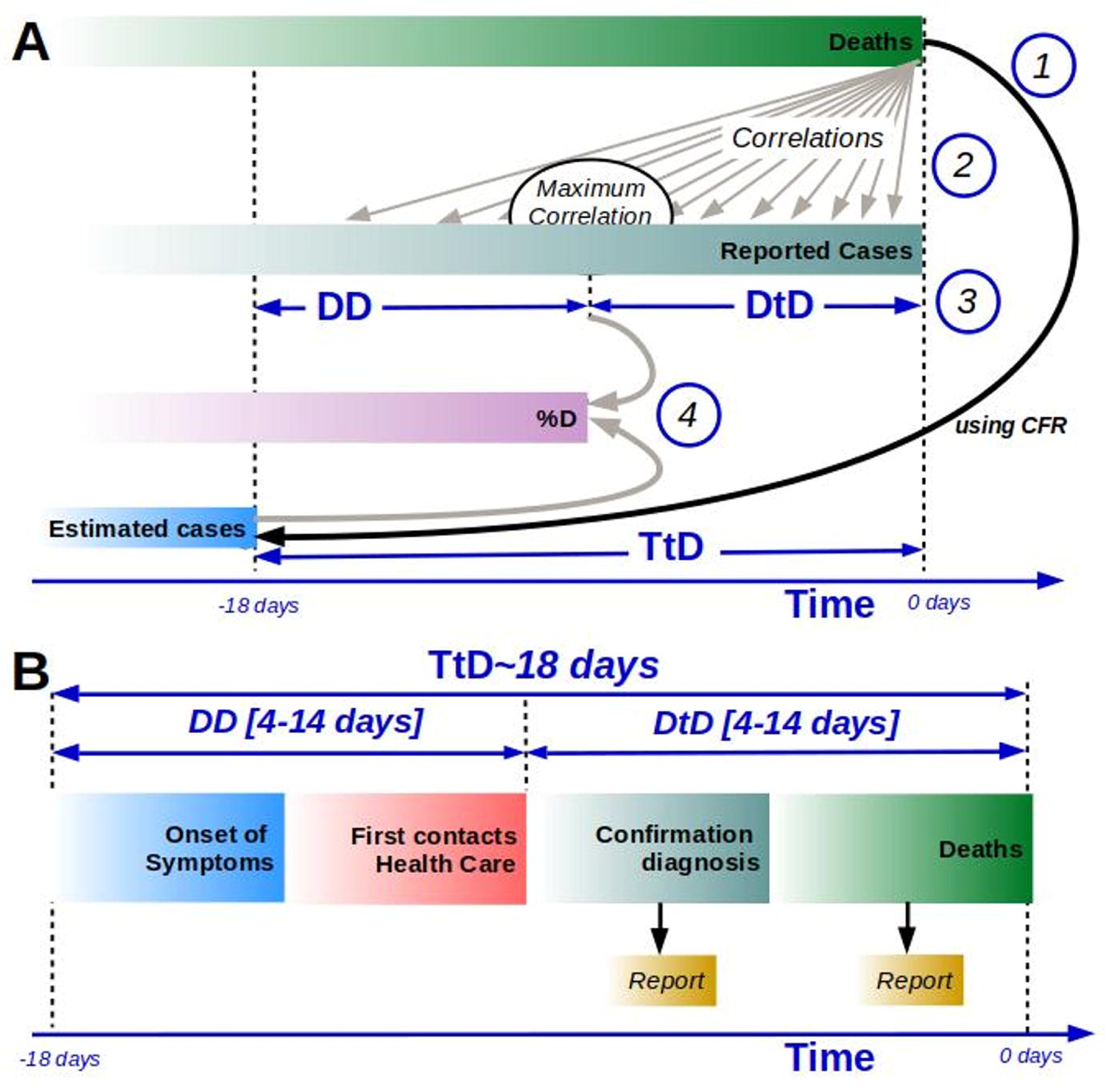
General framework of the calculation of the percentage of diagnosis. (A) Processes involved on the calculation of the percentage of diagnosis: 1.-Evaluation of the estimated cases using TtD and CFR, 2.- Calculation of time correlation between reported cases and reported Deaths, 3.- Evaluation of the time between diagnosis and death (Diagnosis to Death DtD) by the maximum of correlation (country dependent), with DD as the Delay to Detection 4.- Evaluation of percentage of diagnosis based on estimates cases and reported cases. (B) Standard evolution of casualties by COVID-19, from the onset of the symptoms to death, times to reported cases and deaths are shown. Time-lines in the figure are not proportional to real time delays.

This allows us to know to estimate the number of cases 18 days ago. This value can be compared with the number of cases detected 18 days ago, obtaining a diagnostic percentage. This result is an unrealistic lower bound because no one performs PCR testing the first day symptoms, but they are usually done much later. Actually, people normally do not call in a doctor at the first notice of a symptom. Furthermore, depending on the availability of tests, saturation of the health system and other external factors, countries have a great variability in the time of diagnosis delay.

Countries accumulate some delay that may arrive to 18 days in the case that a country detected people as late as they were detected on death. This delay to detection (DD) due to lags in diagnosis corresponds to the time between the patient having the first symptoms and being reported by the health system. In fact, this time in some countries may vary throughout the course of the infection. Therefore we cannot assume that the estimated and the reported are comparable and we need to know what the diagnostic time was for each of the countries studied.

We can compare the reported deaths with the reported cases to find the maximal correlation, see process 2 in Fig. 1(A), to estimate the DD, see process 3 in Fig. 1(A). Finally the ratio between reported cases at DD with the estimated cases, see below, provides an estimation for the percentage of diagnosis, see process 4 in Fig. 1(A). Note that the usual development of the reporting of a new case/death, see Fig. 1(B), depends on the particular country under consideration, which determines DD. In fact, DD also includes a delay in reporting the diagnostic to death to official information systems.

### Real CFR of COVID-19 in Europe

The cornerstone of our analysis is that the real CFR in all European countries will not be biased against any country in particular. We should point out immediately that we are not arguing that there are not important uncertainties in the real CFR, what we do claim and check in this methodology is that these uncertainties will not generate any biased against particular countries and should not affect policy decision. We take the CFR in of COVID-19 in Europe to be between 0.3-3% and we assume 1% to be the benchmark scenario.

This value (1%) is the CFR observed in the initial stages of the South Korea pandemic and the Diamond Princess cruise. In both cases, it was found to be around 1-2.6% and, in both, error margins came from different sources [14, 15]. In South Korea, the ability to test all the population in contact with infected people and the tracking of contagious chains was thorough, despite that, the reported CFR increased from initial values around 0.5-0.7% to higher values around 2%. In the Diamond Princess cruise, CFR for confirmed cases was 2% but estimation of false negatives and the possibility that a fraction of the passengers never developed symptoms and was never tested put the CFR again around 1%. Both South Korea and the Diamond Princess cruise provide complementary evidence, one coming from a natural experiment and another from a country with the ability to perform half a million tests/day from the very beginning of the transmission chain [16]. If we accept the two measurements of the CFR independent, the most likely interval of real CFR is between 0.5 and 2%.

Recent experimental results from random testing in the German city of Gangelt [17] and preliminary results from Iceland [18, 19] indicate the presence of a layer of people fully asymptomatic that are normally not detected. This group of people have passed the disease without any knowledge seems to be larger than previously thought. These preliminary studies point to a CFR of around 0.5% in zones where the epidemics was not fully spread. We cannot disregard the possibility that, just as CFR inceased with time even in South Korea, similar studies in countries with more cases, could have a real higher CFR.

It is thus reasonable to consider CFR at 1% as an easy policy guiding principle and not to use the more positive scenario of 0.5%.

### Unbiased nature of CFR in Europe

There are three sources of possible biased CFR across countries. The disease affects more strongly elder people with comorbidity problems than healthy younger ones, and more men than women. In all European countries the male/female ratio is unbiased except for older people. This is precisely the group with higher mortality rate. It is thus very important to asses how the different demographic structure of European countries could affect our central benchmark [20]. The same must be said about the relative prevalence of other comorbidity factors. We proceed to show that, with the data we have today: the demographic and comorbidity structure, none of these possible sources of bias can have anything but a small effect. To do so, we will do a comparison with the CFR of South Korea on April 15 2020, 2.1%.

Table 1 shows the demographic structure of South Korea and the corresponding CFR for each analyzed age group reported on April 15 2020. The first row shows the demographic structure according to Eurostat, but the analysis has be performed by using only the three age groups shown in the second row: ≤ 49, 50 − 79 and ≥ 80 years. This was done because for many countries reported cases and fatalities consider different age groups, and some countries even report this two figures using different age groups. The three age groups considered in the analysis were the only ones that includes all the analyzed countries. As can be observed in the table, by comparing the percentage of population and cases for the three age groups we can conclude than in South Korea people below 49 years old are infected/detected below their corresponding population importance. The contrary occurs for the people above 50 years old. People above 80 present an increase in infection of 32% with respect their population importance. This is probably due because this people present increased symptoms and they are tested more often.

**Table 1.**
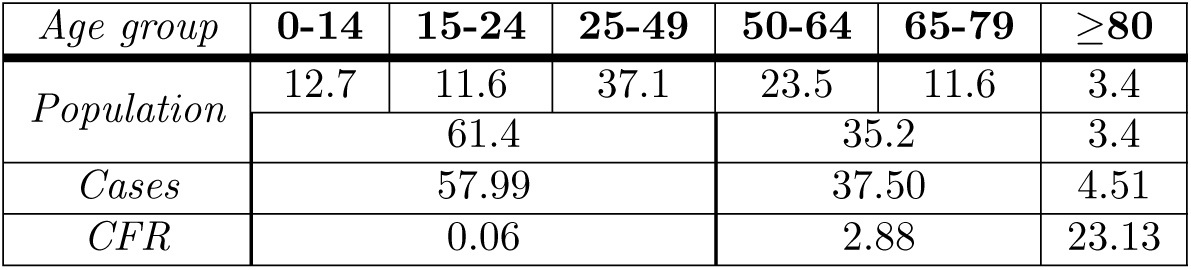
Percentage of population by age group 2016 (median age 41.4), COVID-19 cases and COVID-19 case fatality rate reported in South Korea on April 15 2020. Population age groups are defined following Eurostat criteria. Source: Korea Centers for Disease Control and Prevention, Korean Statistical Information Service.

To analyze what is the role played by the differences in demography in Europe in the COVID-19 cases and fatalities we have downloaded from Eurostat the demographic distribution by age (see Table 2).

We can readily asses that, when comparing with South Korea, all the countries have a larger percentage of population above 80 years (90% larger for Italy) and larger median age except Sweden and United Kingdom, but the relative differences in each of the cohorts in between the European countries shown in the table is small. Only Italy presents a relevant larger than average ratio of people over 80.

Using this demographic data and assuming each European country presents the same CFR by age group as South Korea on April 15 2020, we have computed the CFR for each country. Table 3 shows the results of this analysis and the officially reported CFR by the different European countries on the same date. Both values are presented relative to the CFR reported by South Korea on April 15 2020, 2.16%.

As can be observed in the first column, when demography is the only difference between countries, between the worst and best case of the relative CFR the differences are around 30%. Most countries are in the range between 1.15 and 1.4%, being the average of the relative CFR 1.3%. Therefore, CFR for all the countries is, at most 20% from the average value and typically around 10%.

**Table 2.** Percentage of population by the age groups considered and median age for some European countries. Source: Eurostat 2019.

| *Country/Age group* | $\leq 49$ | 50-79 | $\geq 80$ | Median |
| --- | --- | --- | --- | --- |
| *Belgium* | 61.0 | 33.4 | 4.8 | 41.6 |
| *France* | 60.7 | 33.2 | 6.1 | 41.8 |
| *Germany* | 55.0 | 37.9 | 6.5 | 45.8 |
| *Italy* | 55.4 | 37.6 | 7.2 | **46.7** |
| *Netherlands* | 60.0 | 35.5 | 4.6 | 42.6 |
| *Portugal* | 57.2 | 36.3 | 6.4 | 45.2 |
| *Spain* | 60.5 | 35.2 | 6.1 | 44.0 |
| *Sweden* | 62.0 | 34.0 | 5.1 | 40.5 |
| *Switzerland* | 60.6 | 32.9 | 5.2 | 42.5 |
| *United Kingdom* | 62.5 | 34.2 | 5.0 | 40.2 |
| *EU-27* | 59.1 | 35.1 | 5.8 | 43.7 |

**Table 3.**
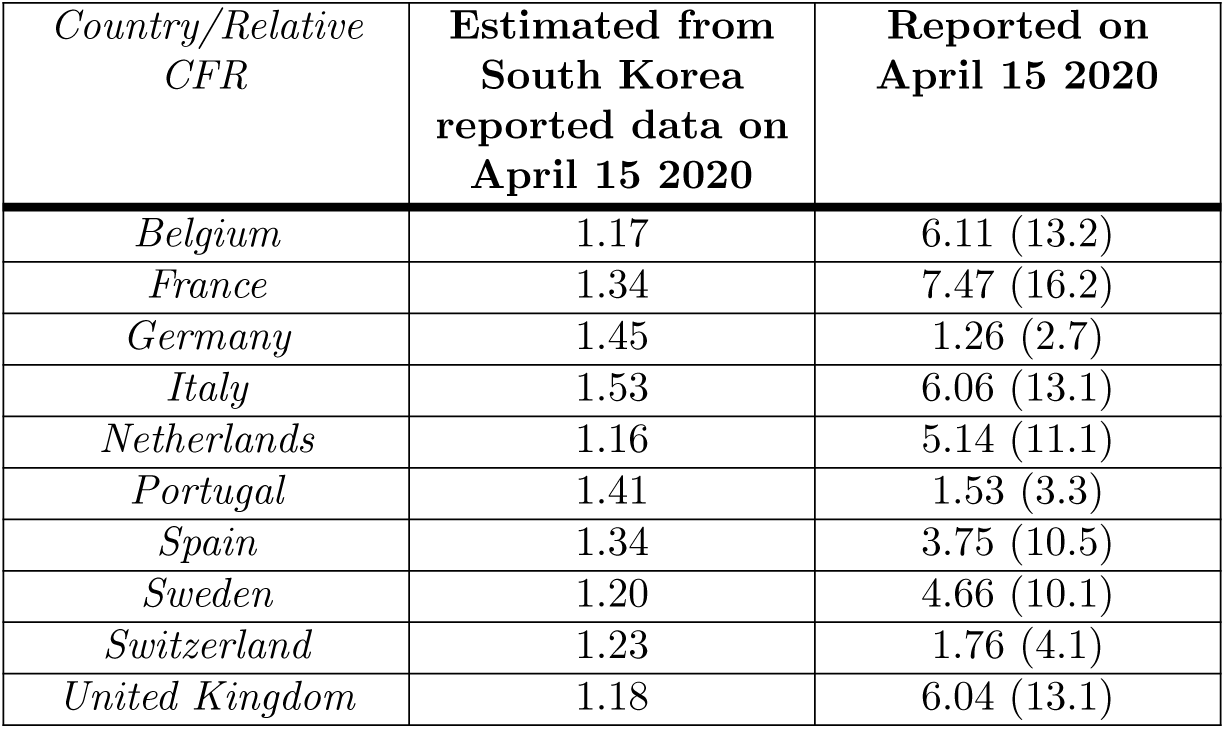
Estimated relative CFR assuming these countries have the same CFR by age group as reported by South Korea on April 15 2020 (see table 2), and officially reported relative CFR on that date. CFR of South Korea on 15 April 2020 was 2.16%. The reported CFR for each country is indicated in parentheses. Source: European Center for Disease Prevention and Control.

### Analysis of possible bias in counting patients deceased due to COVID-19

A unrelated source of bias in the estimation of the real COVID-19 cases, is the possibility that different countries treat and count differently the population that die having previously a very bad prognosis. We know this group is strongly affected by the virus [21]. In blunt terms, we must examine the possibility that different countries are counting the raw number of dead people differently.

Before entering in the detail of the analysis, let us point out that two indications go against this possibility. First, Health Care systems in Europe can have different resources in different countries with different focus and priorities, but they attend anyone with COVID-19 with the exemption of possible patients with multifactorial problems who might be in very fragile conditions. Elder people affected in nursing homes or elder residences who die under suspicious situations are uniformly not reported following European Center for Disease Prevention and Control (ECDC) advise. There is a single exception that we know of: Belgium [22]. Belgium seems to be reporting unconfirmed cases from nursing homes without tests as due to COVID-19. It is quite clear that this includes a good number of people who, either, did not die from COVID-19 or that COVID-19 was not an important factor in the prognosis. Therefore, we will include a reminder that Belgium data is biased compared with other countries, being anywhere from 20% to 50% lower given the number of reported deaths from nursing homes compared with hospitals. There is a second argument regarding the treatment of the elder population in other countries. If large undercounting woul be the case, it should be noted in the mortality rate for people 80 years and older, which is not observed in the countries where we have data.

In this framework, Spain becomes a key country. If Spain were not to have an important undercounting is highly implausible to think that other countries would. We proceed to analyze the data of The National Epidemiology Center (Instituto de Salud Carlos III) of Spain. The center recently published the results of the Daily Mortality Monitoring System (MoMo) for April 18 2020 [23]. They evaluate which periods have mortality well above the average of previous years. When evaluating the period from March 17 to April 18 2020 for the whole of Spain, they see that, as expected, mortality is much higher than in previous years. An increase of 68% is observed. However, it is interesting to compare this with the data reported for COVID-19 deaths. The reported deaths by COVID-19 are roughly 20000, depending on how you attribute deaths to a particular day in the calendar. On the other hand, the reported excess of deaths by the MoMo surveillance system is 25000. We think that the assessment of around 20% underreporting can be taken indeed as a worst-case scenario for a highly impacted country. It seems reasonable to expect other countries to have underreported way below or slightly below this level. All the data point out right now, that the undercounting due to a different treatment of the very fragile population is highly unlikely across Europe, and at most introduces changes in CFR around ±10%.

### Treatment of cross-country bias in diagnostic time-delays

Having shown that the real CFR should not present bias in European countries larger than 25%, we address now how to deal with the real sources of bias in the diagnostic rate for each country. To estimate DD we look for a correlation between the number of reported cases (see Fig. 2A) and the number of reported deaths (see Fig. 2B) [1, 24]. To deal with noise effects we put a weighted moving average filter on the data of both cases and deaths. The correlation time between reported cases and reported deaths will be named as time from diagnosis to death (DtD), and:

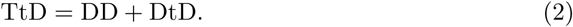

**Fig 2.**
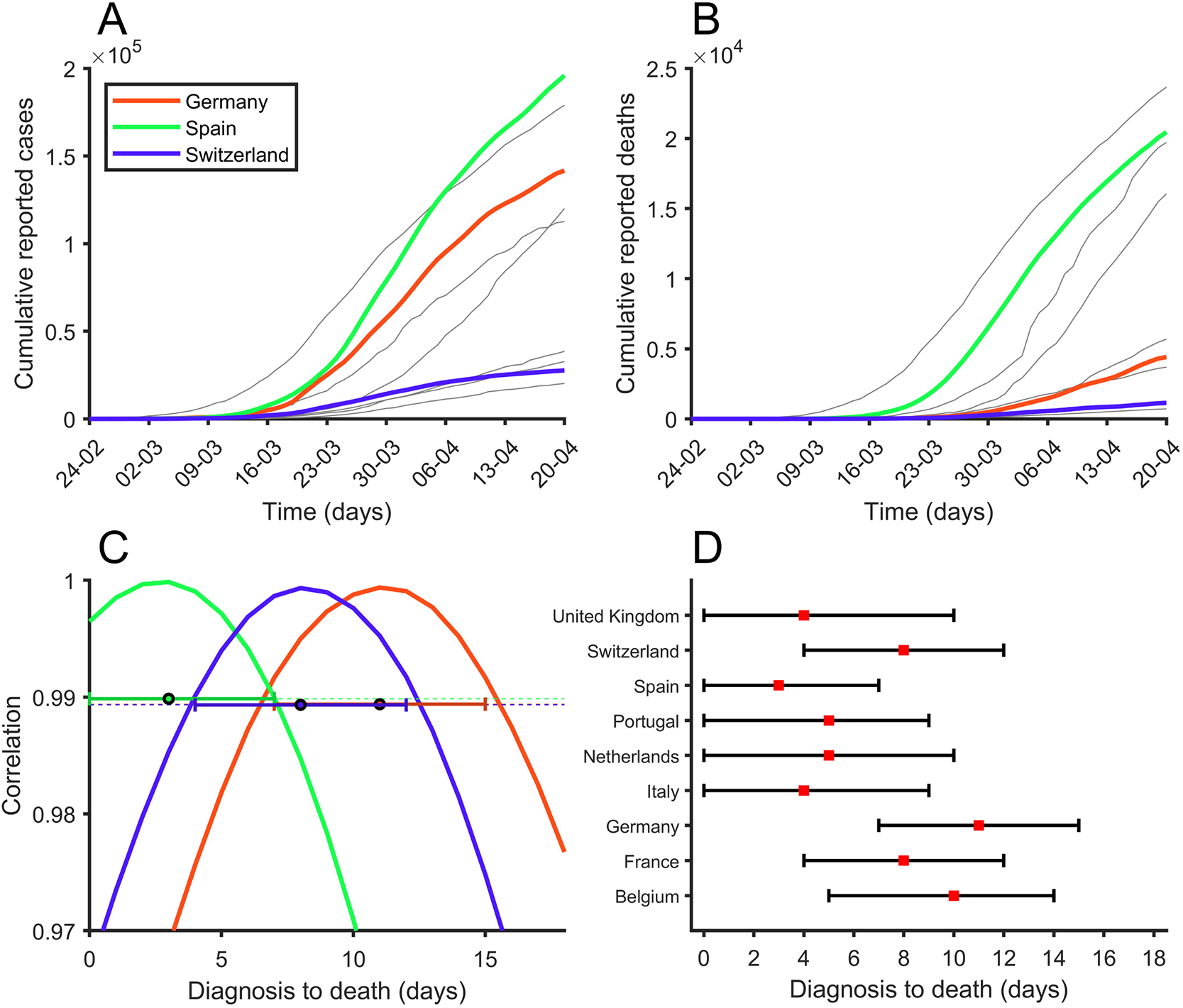
Correlation between reported cases and deaths. (A) Number of cumulative reported cases, (B) Number of cumulative reported deaths and (C) Correlation between reported cumulative cases and reported cumulative deaths exploring different delays between diagnose (reported) and death for Germany (red), Spain (green) and Switzerland (green). (D) Maximum correlation is marked with a red square for each country. 99% correlation interval can be seen with black bars.

In Fig. 2C we can see the correlation [25] between reported cases and reported deaths assuming different DtD for Germany, Spain and Switzerland. As you might expect, correlations have values close to 1. In most cases the correlation has a concave parabolic shape with a clearly defined maximum. We assume this maximum represents DtD for each country. The correlation interval is estimated as the points where the correlation is greater than 99% of the observed maximum. We decided to set a lower limit of 4 days and a higher limit of 14 days [11] because we believe that time outside this range would be unrealistic situations. For countries that have not seen a clear correlation (Sweden) it has been decided to explore the entire DtD interval (4 to 14 days). Approximate values for DtD are shown in Fig. 2D. In the supplementary material the correlation curves can be observed for the 10 analyzed countries. Then the percentage of diagnosis cases diagnosed at time reads:

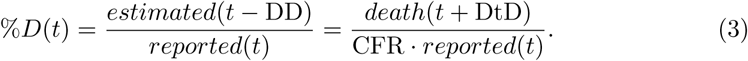

## Results

### Diagnostic rate by country

As discussed in the methods, we use the same CFR = 1% in all European countries instead of making small corrections for demography. The bias due to demography was shown to be around 10-15%, precisely the same order of magnitude we obtain for the possible bias in the counting of reported mortal cases. Given that our aim is to provide a clear method for policymakers and that there is no data on how, or even if, both correlate, a common CFR allows us to homogenize the results with the clear limitation that we will obtain reasonable estimations and not exact results. The resulting picture is expected to be closer to reality than using purely reported data, but worse than correcting properly for age and diagnosis if the data of CFR for all age brackets and locations (nursery homes, hospitals, individual homes) were available, which is not the case.

The estimation of the diagnostic rate is straightforward. From the cumulative number of deceased each day, and multiplying by 100 (1% CFR) we get the cumulative number of people with symptoms 18 days ago [10–12] simply by rescaling and displacing backward in time the cumulated death curve of any country. To give an initial realistic and homogenous diagnostic rate we must establish how many days are needed as a bare minimum to be able to detect a patient from the onset of symptoms. First, the patient has to feel sufficiently sick and then contact the Health Service. From this contact, the doctor needs to be suspicious that the person has the disease and request a test. Then, this test must be available, performed and the result received and annotated. It is clear that a bare minimum of one week is needed for this process. We use the name 7-Days Diagnostic Rate (7D-DR) for the diagnostic rate with a benchmark of one week of diagnosis delay.

We explain the procedure to get 7D-DR for each country in the top left panel of Figure 3. We take the cumulated death curve of any country, rescale according to the CFR and displace it towards the past 11 days. This is 18 days back to the onset of symptoms and then 7 days forward to be detectable/diagnosable. From this curve, we can obtain the rate between the cumulated number of people who had symptoms for 7 or more days and the cumulated number of people detected 11 days ago. It is thus clear that this homogenous analysis across countries could be performed assuming 5D-DR or 9D-DR and different CFR. It gives a proper first estimation of the situation.

**Fig 3.**
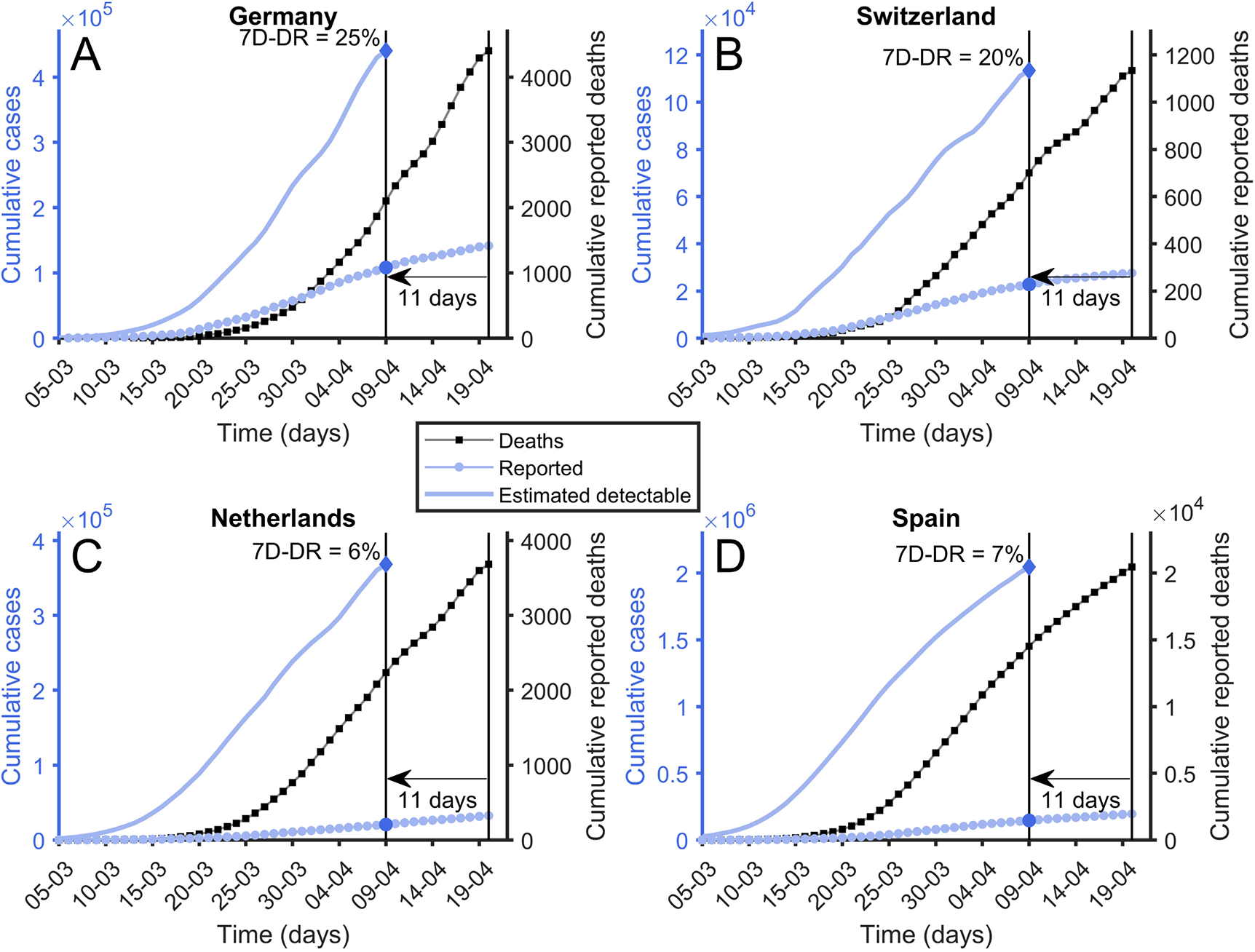
7-Days Diagnostic rate. Reported cumulative number of deaths (black squares), reported cumulative number of cases (blue circles) and estimated number of cases calculated using Eq. 1 (solid blue line). To compute 7-Days Diagnostic Rate it is used a diagnostic to death time of 11 days. Its value is calculated using last available points. (A) Germany. (B) Switzerland. (C) Netherlands. (D) Spain.

We argue, however, that there is indeed bias in the way people deal with the health care system in normal situations and, especially, under an epidemic. Different countries and populations are in fact behaving very differently. We have observed that this is the case in the methods section checking the delay between diagnostic and death using time-displaced correlation analysis. This is the reason why we also define the Delay to Detection Diagnostic Rate (DD-DR) as the diagnostic rate computed using a time delay between the appearance of symptoms and detectability different for each country. We proceed to use Fig. 4, with Spain as an example, to explain the concept behind DD-DR.

**Fig 4.**
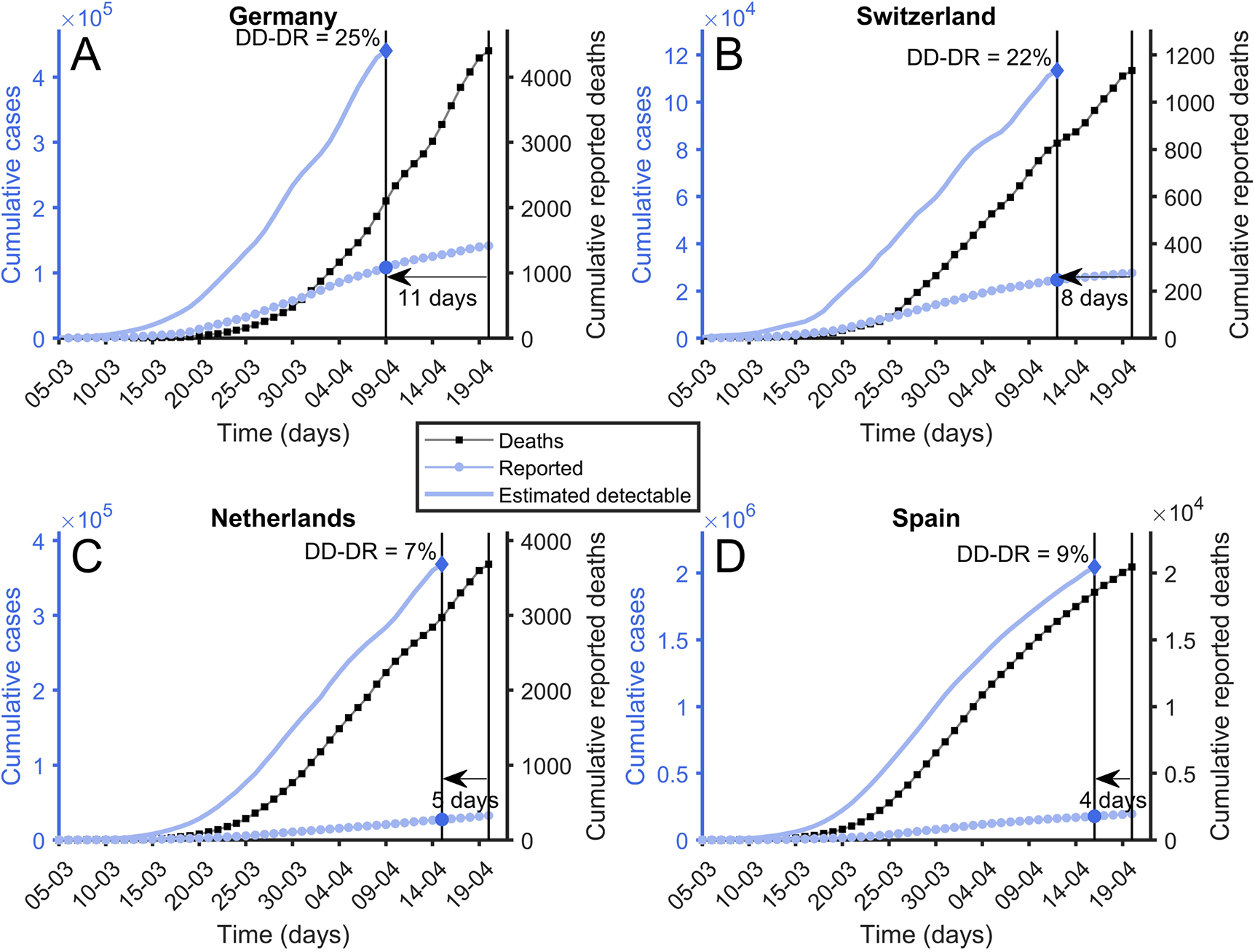
Delay to Detection Diagnostic Rate (DD-DR). Reported cumulative number of deaths (black squares), reported cumulative number of cases (blue diamonds) and estimated number of cases calculated using eq 1 (solid blue line). To compute Delay to Detection Diagnostic Rate it is used the diagnostic to death time observed in Fig 2. Its value is calculated using last available points. (A) Germany, DtD = 10 days. (B) Switzerland, DtD = 8 days. (C) Netherlands, DtD = 5 days. (D) Spain, DtD = 4 days.

For Spain, the maximum correlation between cumulated death curves and cumulated reported cases appears when cumulated deaths are displaced 4 days backward. This suggests a DD of around two weeks (18 − 4 = 14 days). This makes sense in a situation like the one in Spain during March 2020. The population receiving news that the health care system is under stress may decide to delay reporting of symptoms unless they are very serious. Additionally, there is the possibility that tests are not available to people who report with symptoms to primary health care centers, and that the delay between the test, its positive result, and its record to official information systems is not negligible as well.

It is thus important to correct for this bias in the estimation of the diagnostic rate. It is clearly not the same to have a time delay from symptom to the detection of 14 days than 7. DD-DR can be computed from Spain just like we did before for the 7D-DR using the same rescaling of the cumulated dead curve as before but using a displacement backward of 4 days instead of 11 days. Fig. 4 shows how the DD-DR is obtained in different countries depending on the delay between symptoms and detectability. Countries with a lower DD, such as Germany, have the same 7D-DR than DD precisely because they diagnose as early as realistically possible.

We notice now that both 7D-DR and DD-DR can be tracked along time, as the epidemic advances we can check how these diagnostic rates changes. Each new day we can look 11 days back for the 7D-DR and compute the diagnostic rate. DD-DR can be tracked similarly. In Fig. 5 we show the evolution for both as a function of time for three selected countries. We observe that the DD-DR reaches a steady state after the initial stages of the disease while 7D-DR seems more affected by trends. This can be expected since DD-DR uses, precisely, the maximum of the correlation delay so it is expected to fluctuate less. The DD-DR is not only more stable but it also allows as to produce a proper assessment of the errors involved. The main one is the fact that the exact delay between onset of symptom and detectability in each country has large uncertainty. While the best estimation of the time delay in Spain is 14 days, the real value could be also around 12. For Germany, for example, DD can be anywhere from just 4 days to 11 days. Using different time delays we obtain different diagnostic rates. It can be observed in Fig. 5 that the percentage of diagnoses of Germany or Switzerland is approximately constant over time. In the Supplementary Information, we show the evolution of DD-DR for the 10 countries studied SI Fig. 1. Assuming that this percentage remains constant to this day and that the diagnostic conditions have not changed over the last few days, we can estimate the total number of cases as of mid April 2020 as above 2 million in Spain and close to half million in Germany.

**Fig 5.**
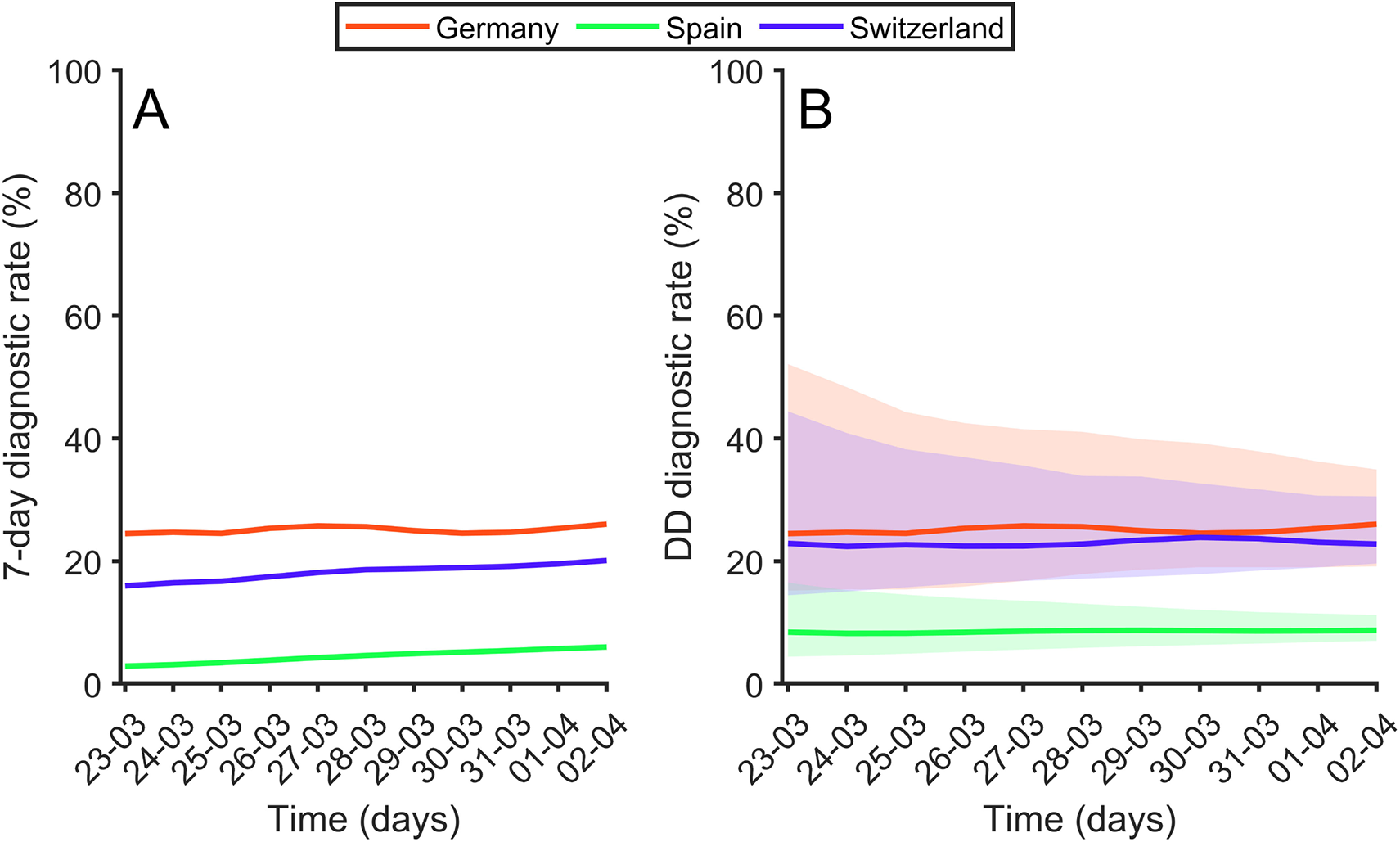
Diagnostic rate along time. (A) 7-Days Diagnostic rate along time for Germany (red), Spain (green) and Switzerland (blue). (B) Delay to Detection Diagnostic Rate along time. Thick lines are using the Diagnostic to Death time observed in Fig 2. Shaded parts represent the limits considering error bars observed in Fig 2.

The table in Fig. 6 shows a list of the 7D-DR as of the beginning of mid-April of 2020, and the DD-DR, which seems stable, together with the associated error.

**Fig 6.**
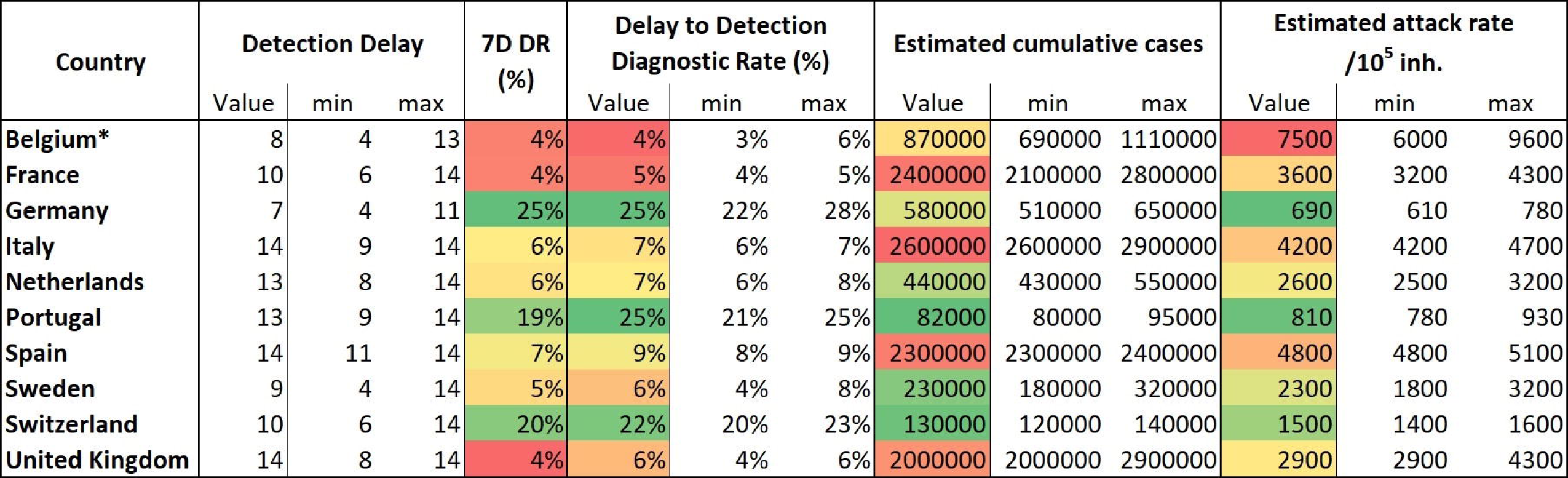
Detection delay (DD), 7-Day Detection rate (7D-DR), Delay to Detection Diagnostic Rate (DD-DR), estimated cumulative cases and estimated attack rate. To interpret estimated cumulative cases and estimated attack rate we must take into account Detection Delay, because they are computed using the reported data. Data updated on April 20 2020. Belgium data is biased due to reporting of unconfirmed death cases [22]. Best estimations might shift 20-50 %

### Effective growth potential (EPG) index for policy makers

Once the diagnostic rate is known, it is straightforward to establish a real incidence no longer affected by the presence of important differences in the time delays to diagnostic in different countries (see the table in Fig. 6). The level of diagnosis and the real incidence is indeed useful for policymakers since it gives a clear general picture. However, the policy response needed to improve the diagnostic rate is limited, in the short-term, by the ability to increase the production of PCR kits and other diagnostic tools.

Policymakers have more ability to affect immediately mobility patterns and social contact. In this sense, a key number for policymakers would be to have a reliable and robust estimation of the number of infected people in each country that can propagate the disease. Providing an exact number is, right now, impossible.

We can, however, produce an index of the effective potential growth using the DD-DR and the guidelines used by the ECDC to track the epidemic. Even if the precise number of people with the disease were known, and the distribution of symptoms by sex and age was reported, there is no clear knowledge regarding the level of infectivity of the different type of person and symptoms. For instance, it is not known the days a person with mild symptoms can transmit the disease. The same can be said for people with serious symptoms. Virus loads in the throat seem to be rather high across the board [26], but data on how this influence contagion is unclear. The only way to assess the situation is to use a general unbiased broad measure, which is indicative of the potential for infection. The ECDC uses the number of newly infected people in the last 14 days [27]. We use this same criterion.

Figure 7 shows how to compute an estimation of the people that go undetected and have the potential to transmit the disease. Using the DD-DR one can compute how many undetected people were added to the infected number in the last 14 days, I_14_. This number can only be obtained properly some days in the past, on the day we have a typical diagnosis. After that, we would need input from new data to properly compute how many people are diagnosed. So the number I_14_ is strictly a measure of the recent past, but good enough to give the proper picture that the system will face the following days.

**Fig 7.**
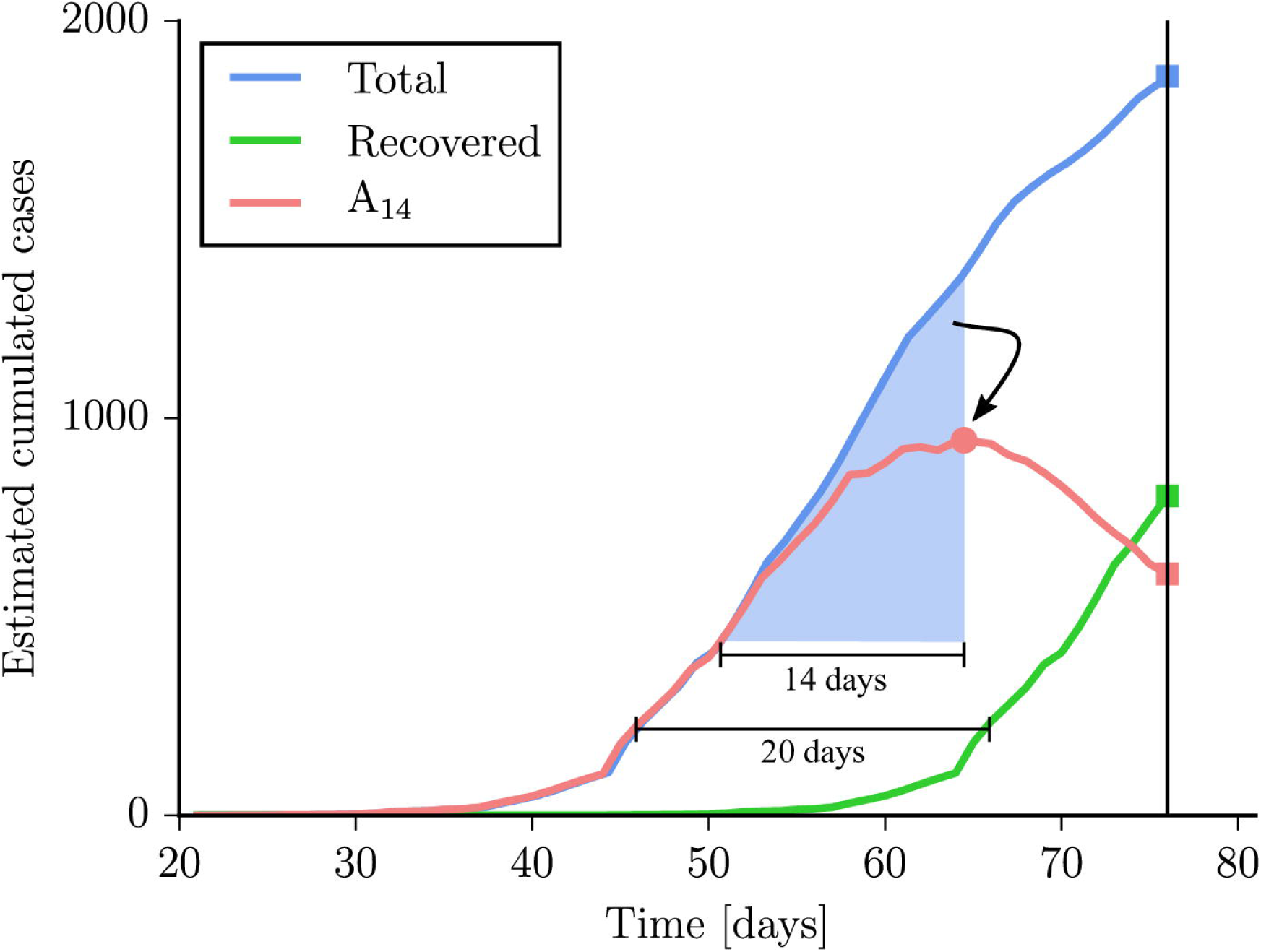
Schematics of the procedure to obtain incidence A_14_, recovered and estimated cases using Germany as an example. Incidence of estimated cases (blue), contagious incidence (red) and total estimated recovered cases (green). Blue shaded part is the number of cases used to compute the estimated contagious incidence. To interpret final number of total cumulative cases, recovered cumulative cases and estimated attack rate we must take into account Detection Delay, because they are computed using the reported data. Similar figures for all countries are shown in SI Fig. 2

We also consider those undetected cases which appear earlier than 14 days as recovered *R_I_*. Notice that here we use the word recovered lousily. It does not mean literally that all of them are fully recovered since most of them never fell ill to begin with, and some of them could not have neutralized tests yet, but that those infected and undetected for more than two weeks ago do not seem to pose a serious risk.

A list of values for I_14_ and the corresponding 14-day attack rate per 10^5^ inhabitants (A_14_) are provided in each country in the table of Fig. 8 with the number computed at the beginning of April 2020. These values are currently been monitored each day for all UE countries. Having an unbiased assessment of the risk regarding the number of potential spreaders, I_14_ and A_14_, are key values for policymakers.

**Fig 8.**
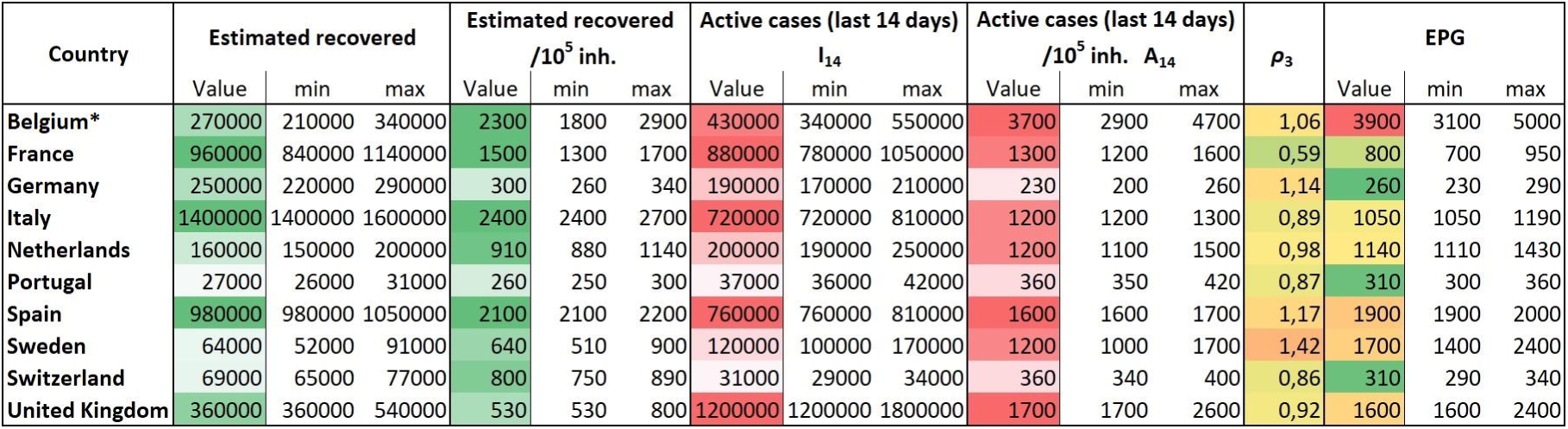
Estimated recovered and active cases, 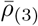 and EPG. 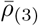 is computed using the mean value for the last three days. EPG: Effective Potential Growth described in the text. To interpret table data we must take into account Detection Delay, because they are computed using the reported data. Data updated on April 20 2020. * Belgium data is biased due to reporting of unconfirmed death cases [22]. Best estimations might shift 20-50 %.

**Fig 9.**
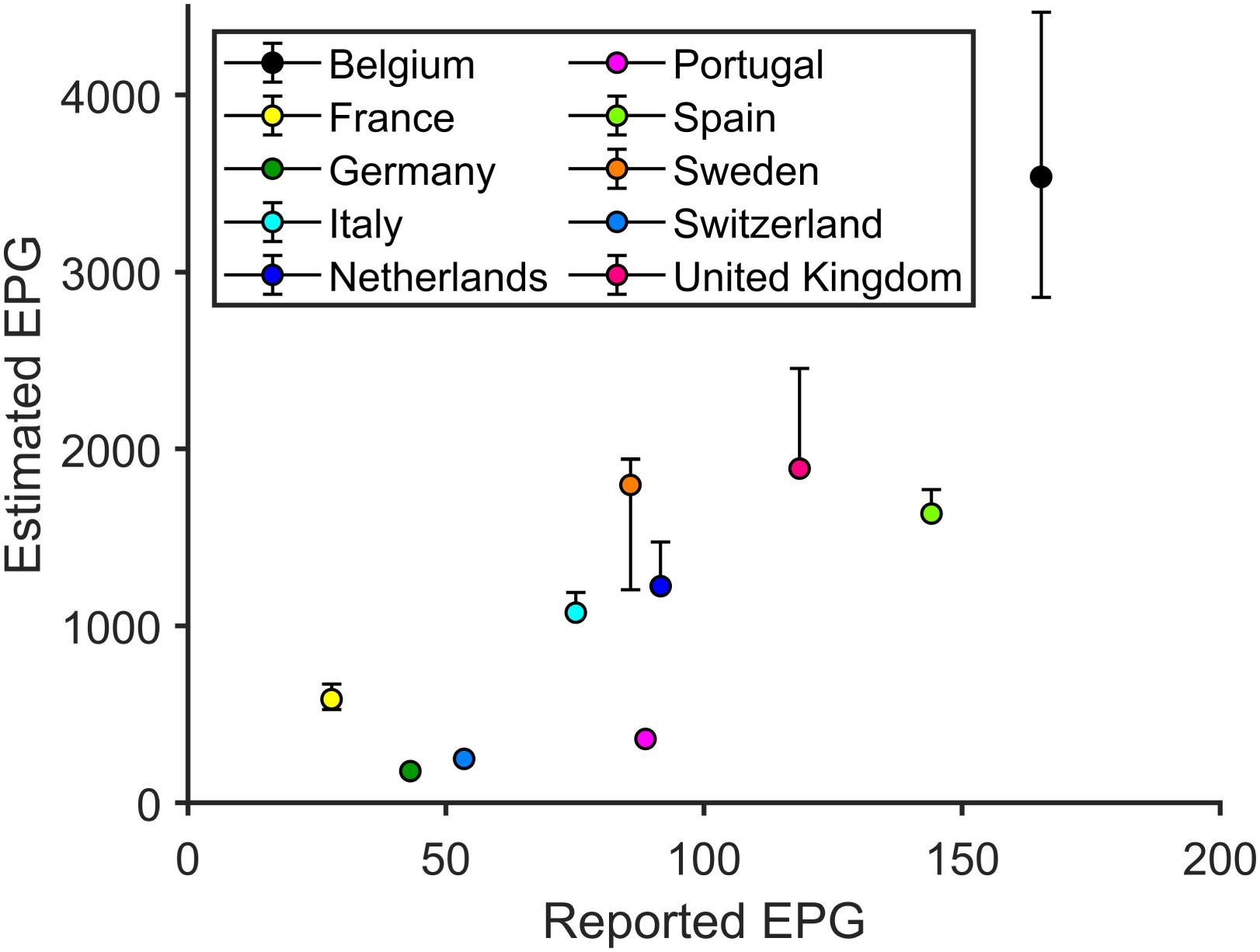
Reported EPG vs estimated real EPG. Different European countries in terms of the EPG computed using the reported data on the attack rate vs the EPG using our estimation of the real attach rate. The order of the different countries should be done from right to left (for the reported state of the index) and from top to bottom (for the estimated value of the index). We observe how the comparative situation of the different countries changes as of 20 April 2020. * Belgium data is biased due to reporting of unconfirmed death cases [22]. Best estimations might shift 20-50 %.

A_14_ alone, however, does not give a full picture of the situation. It is not the same to have 100 contagious per 10^5^ inhabitants when the number of contacts is high that when the number of contacts is low. It is important to take into account the level of spreading velocity of the epidemic related to the effective reproductive number (*R_t_*).

The effective reproductive number depends on multiple factors, from the properties of the virus itself to the number and type of contacts. Those, again, depend on different social behavior and structure such as mobility, density or the typical size of the family unit sharing a house, to name a few. The only feasible way to estimate *R_t_* is using fits from SEIR models. Complex SEIR models which include spatial and contact-processes have a large number of parameters which, due to the present lack of knowledge, are unknown. This makes any estimation of *R_t_* highly dependent on the value of other co-factors who affect strongly the propagation. Effectively, *R_t_* can be only fit in very simple SEIR models where a small number of parameters are unknown and *R_t_* can be calibrated from. This makes those models basically empirical.

Given the partial empiric nature of present *R_t_*, we prefer to take a fully empiric surrogate as a quantitative evaluation of the level of infections. We define an alternative reproductive number as the number of new cases detected today divided with the number of new cases detected five days ago as *N_t_*/*N_t−_*_5_. However, the high fluctuations on this quantities imposes the use of averaged values over three days [9]:

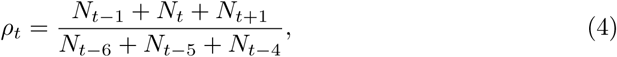

where *N_t_* stand for new cases reported at day *t*. This rate is one if the number of new cases is constant. It will be below 1 if new cases are decreasing and larger than 1 if the number of cases is increasing. We take 5 days as the key delay unit since this is roughly the time since infected people develop symptoms if they do develop them.

There are still clear fluctuations on a day-to-day basis of this measure *ρ_t_* due to common delay and irregularities in reporting. Most fluctuations can be eliminated by taking the average of *ρ_t_* during three days 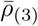

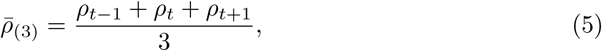

which is normally enough to get a rather smooth measure. It is not uncommon to find still some fluctuations and one-week averages can be done if required.

We propose the following day-to-day index EPG:

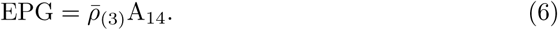

EPG is just the multiplication of the growth rate of the disease 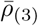 with the estimation of A_14_ both evaluated at the proper time in the recent past. The worst scenario is one where both A_14_ and *ρ* are large. It means you had a lot of population with the disease and lots of spreading a few days ago. The best situation is a low value of velocity and the number of active cases. Having a large number of A_14_ with low 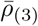 or a large 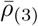 with low A_14_ are potentially dangerous situations. On April 20 2020, these values are given in the table of Fig. 8 for the European countries we tracked. These values can be updated every day [9].

## Discussion

Reported number of deaths per 100,000 population is a fairly objective and relatively simple way of assessing the situation of COVID-19 epidemic in countries. The complete picture must be given by a more complex analysis of other data such as the number of diagnoses per 100,000 inhabitants, distribution of these cases among regions and according to age and sex, percentages of asymptomatic and mild cases, and spreading rate of the epidemics, among others. Nevertheless, any analysis based on diagnosed cases is biased by diagnosis protocols and ratios in each country, as well as by the pool of asymptomatic cases. Moreover, any attempt to improve diagnosis percentage requires an economic, infrastructural and logistical effort that is not always possible. In addition, this health system structure is a strong conditioning that limits the possible actions to carry out in this direction. The reported number of deaths, if uniformly and properly recorded, provides very relevant information as a first general overview. Even in countries where there is a bias on death reporting, the effort that should be made to improve these data collection is much lower than the necessary effort to increase data about cases.

The assumption of a common lethality, which has been situated around 1%, allows for using the CFR as an indicator of real incidence. Current information on CFR is still not complete, since many countries do not report distribution of deaths by age or sex, neither provide COVID-19 mortality outside hospitals. However, we argue that the picture that we obtain from the analysis of CFR is closer to reality than the one provided by the pure analysis of reported cases. In particular, this analysis allows for (1) establishing an order of magnitude of real cases and diagnosis percentage, (2) assessing an effective potential growth index that evaluates the risk, and (3) obtaining an order of magnitude of recovered people that could be potentially immunized at short-term.

In Europe, absolute cases’ ranking has been lead by Italy (until April 4 2020) and Spain (since then). On April 20 2020, Spain was at the level of 196,000 reported cases while Italy was reporting 179,000. They were followed by Germany (142,000), United Kingdom (120,000), France (113,000) and Belgium (38,000). If we estimate the cases that should have been diagnosed by that time, the ranking is lead by Italy (2,600,000) and followed by France (2,400,000), Spain (2,300,000), United Kingdom (2,000,000), Belgium* [22] (870,000) and Germany (580,000). Thus, differences in diagnostic rate are absolutely significant when analyzing global situation in Europe. Countries like Germany, Portugal and Switzerland would be diagnosing around 25% of cases, while Belgium, France, Sweden and United Kingdom would be in the level of 5%.

Assessing the risk of countries to enter or remain in the epidemic growth phase is essential. In this sense, the EPG index is a valuable tool for policy makers. A high EPG in the situation where there is a high growth rate of the epidemic and large number of active cases is a clear situation of danger, while a very low EPG because both the number of recently infected and the spread velocity are low is a clear situation of control. In intermediate situations, EPG informs whether the growth rate is too high for the number of infected at hand. Even if 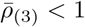 and the epidemic seems to be under control because new cases are decreasing, intermediate EPG informs the policy maker that reopening can have a very important cost in the form of secondary focus and waves of infection. A rather large EPG with low 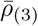 is a situation where the number of spreaders is potentially very high and increasing the number of contacts carries a large risk. Therefore, EPG is very informative index that also is very robust.

Despite 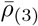 is quite independent of the diagnostic rate, reported I_14_ directly depends on the level of diagnosis. Thus, if EPG is evaluated with reported data, it can provide a wrong picture of the situation. Based on reported EPG, the worst situation in Europe at April 20 2020 would be for Belgium, followed by Spain, United Kingdom, Netherlands and Portugal. If risk is evaluated with estimated EPG, highest value would still correspond to Belgium as well, but followed by Sweden, United Kingdom, Spain, Netherlands and Italy. Portugal is in much better position that its reported data suggest. Actually, countries with similar reported EPG like Portugal, and Netherlands have, in fact, totally different estimated EPG, being the last country at significantly higher risk than the former 9.

We have shown in the Methods section that the basis for obtaining estimated I_14_ and A_14_ is not biased due to demographic differences and, right now, there is no indication that it is biased due to a different way of accounting for the cumulative dead toll of the epidemic. There is also no indication that comorbidity factors are largely different in different countries or that CFR is higher on some countries given that ICU units and hospitals are not available for people that would need it, at least so far. If this were the case, under any scenario where the situation occurs, the epidemic in that country will have such a larger number of cases, attack rate and growth that the EPG will be extremely high. The only real limitation is that the social and environmental issues could affect the prognosis of the infected. If living in a small house with other people infected could lead to worse prognosis than staying in a large house alone, a new analysis regarding the unbiased nature of the CFR would need to be done.

It is important to indicate that not only I_14_ is unbiased, as analysed in previous sections, but that 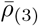 is not biased as well. Even though absolute reported cases is biased, as we have shown, *ρt* deals with ratios and its evolution. As long as the diagnosis and recording of the people with disease follows roughly the same criteria along time in each country, *ρ_t_* is a good measure of the growth the epidemic. Indeed, if evaluated diagnosis percentage is more or less constant in time, we can assume that *ρ_t_* correctly reveals tendencies in contagiousness. If a change in criteria in reporting the cases occurs (i. e., a large increase in the number of tests per day leading to an increase of cases due to more testing), *ρ_t_* will be temporally affected but will go back to be a good measure once the new criteria is established. In this case, EPG will provide a wrong picture for a while as well, until stationary conditions in diagnosing and reporting are achieved again.

There is another important point to address in order to guarantee that *ρ_t_* is a robust measure. As soon as we are estimating real number of cases, we can determine the associated *ρ_t_*. It is expected that both *ρ_t_* behave similarly but with a certain delay. This delay can be determined by translating both *ρ_t_* in time until error between both is minimized. We show this detailed analysis in the Supplement Material SI File where we evaluate that both the reported *ρ_t_* and the inferred *ρ_t_* are indeed different but that follow the same type of evolution once the proper delay is accounted for SI Fig. 3.

The third important outcome of this analysis is the estimation of recovered people. This is an important number to assess the possibility of herd immunity discussed as a possible exit strategy. The idea is that those that recover might have immunity and act as barrier in the transmission of the disease. A recent study from the Fudan University at Shangai [21] has analyzed antibody titters of 175 adult COVID-19 recovered patients. The study is based in the detection in plasma of Spike-binding antibody using RBD, S1, and S2 proteins of SARS-CoV-2 using an ELISA technique. It is also the first study that looks after neutralizing antibodies (NAbs) specific for SARS-CoV-2 using a gold standard to evaluate the efficacy of vaccines against smallpox, polio and influenza viruses.

The study highlights the correlation between the NAb titters and Spike-binding antibodies that were detected in patients from day 10-15 after the onset of the disease, remaining afterwards. Middle and elderly age patients had higher titters compared with young age patients, in which in 10 cases the titters were under the limit of detection. NAb titters had a positive and negative correlation with C-reactive protein (CRP) levels and lymphocyte counts, respectively. This indicates that the severity of the disease, in terms of inflammatory response (CRP levels), usually worse in middle and elderly age, favors the increase of antibody titters. Equally, the negative correlation with lymphocyte counts suggests an association between cellular and humoral response. Therefore, it is possible that the immunity reached by young people, which were mostly asymptomatic, is residual. In that case, this sub-population would keep being carriers of COVID-19. Serological studies that many countries are designing and carrying out should provide further information on post-infection immunity.

Even if the entire recovered population acquires middle-term immunity, current incidence situates European countries far from herd immunity. Nevertheless, it is feasible that regions with highest affectation were closer to use herd immunity as a strategy for de-confinement. Governments might wish to explore the possibility of local deconfinement.

## Limitations

There are two possible limitations of this present study. It could be possible, in theory, that some countries present an intrinsically different CFR if they are able to isolate completely and significantly its elder population more than others. The epidemics real CFR is a measure of the case fatalities if all the population, or a representative sample of it, has become infected. If one country would effectively prevent all infections among all its elder population from contagious forever, it will certainly have a different CFR.

Right now, it is impossible to assess if this is indeed the case in different countries given the lack of reported cases and mortality rates by age and sex. We should notice however that, if this disaggregation were to be provided, we could proceed with exactly the same methodology but instead of using the whole country as a whole we would divide it into different age brackets and treat them separately.

The second limitation is related to the first one but coming from a more structural perspective. A clear possibility is that countries under stress could be failing in providing the same medical support changing the CFR. We must notice that Health Care in European countries, even under stress, has been able to increase dramatically its number of health personnel, of beds and hospitalization in short notice [28, 29]. Italy and Spain present some regions under stress but not the whole country [30]. Finally, one cannot disregard the possibility that complex mechanisms of mutations and repetitive exposure to the virus may change the prognosis depends on the type of residence and, hence on socio-economic factors, which are clearly different across countries. If any proof that a close environment not only increases the level of infections, which they obviously do, but also changes the disease evolution in the patient, one should again test that the uniform/unbiased CFR hypothesis holds with the proper knowledge at hand.

## Conclusion

We have estimated the diagnostic rate of European countries in an unbiased way and reported EPG (Effective Potential Growth) as an effective index to monitor the comparative situation of COVID-19 in different European countries. The diagnostic rate is different in each country but roughly constant in time. On the other hand, EPG changes for each country and at each stage of the epidemic becoming large when there is a worrisome situation.

## Supporting information

**SI File. Supporting Information file**. It includes figures showing the correlation to obtain DtD for each country and the corresponding evolution of the diagnostic rate. We also provide fore each country the evolution of recovered and the attack rate in the last 14 days A_14_. We also provide the demonstration that 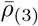 is also unbiased showing the correlations between real and estimated growth rates.

**SI Fig. 1 Series of figures showing diagnosis to death (DtD) for different European Countries**. (A) Correlation between reported number of cumulative cases and reported number of cumulative deaths using different DtD times. (B) Alignment between reported number of cumulative cases and reported number of cumulative deaths using three different detection delays (DD). (C) Diagnostic rate along time using different DD, from top to bottom 0 to 18 days. In red, 7-day detection rate and, in blue, Delay to Detection Diagnostic Rate.

**SI Fig. 2 Series of figures showing the evolution of the estimated cases for different European countries**. In blue, incidence of estimated cumulative cases. In green, estimated incidence of cumulative recovered cases. In red, estimated incidence of attack rate lasts 14 days (A_14_). Day 1 is considered the first day where cumulative cases was over 100 cases, it is different for each country. Data extended till April 20 2020.

**SI Fig. 3 Series of figures showing the relation between reported growth rate and estimated growth rate of the epidemic**. (A) In green, estimated cases growth rate and, in blue, reported cases growth rate. (B) The gorwth rate of estimated cases is displaced to find better match with the growth rate of reported cases. (C) Error between estimated and reported growth rates using differents delays. Minimum delay is marked and is the one used in (B).

## Data Availability

Data is available from ECDC repositories

## Acknowledgments

CP, PJC and MC received funding from La Caixa Foundation (ID 100010434), under agreement LCF/PR/GN17/50300003; PJC received funding from Agència de Gestió d’Ajuts Universitaris i de Recerca (AGAUR), Grup Unitat de Tuberculosi Experimental, 2017-SGR-500; CP, DL, SA, MC received funding from Ministerio de Ciencia, Innovaciòn y Universidades and FEDER, with the project PGC2018-095456-B-I00. EA-L received funding from Spanish Ministerio de Economía, Industria y Competitividad under grant number SAF2017-88019-C3-2-R. This project has been partially funded by the European Comission - DG Communications Networks, Content and Technology through the contract LC-01485746

